# Insights into genetic factors contributing to variability in SARS-CoV-2 susceptibility and COVID-19 disease severity

**DOI:** 10.1101/2021.05.10.21256423

**Authors:** Matteo D’Antonio, The COVID-19 Host Genetics Initiative, Timothy D. Arthur, Jennifer P. Nguyen, Hiroko Matsui, Agnieszka D’Antonio-Chronowska, Kelly A. Frazer

**Affiliations:** Department of Pediatrics, University of California San Diego, La Jolla, CA, 92093, USA; Biomedical Sciences Graduate Program, University of California, San Diego, La Jolla, CA 92093, USA; Department of Biomedical Informatics, University of California, San Diego, La Jolla, CA, 92093, USA; Bioinformatics and Systems Biology Graduate Program, University of California, San Diego, La Jolla, CA, 92093, USA; Institute of Genomic Medicine, University of California San Diego, 9500 Gilman Dr, La Jolla, CA, 92093, USA

## Abstract

Variability in SARS-CoV-2 susceptibility and COVID-19 disease severity between individuals is partly due to genetic factors. Here, we applied colocalization to compare summary statistics for 16 GWASs from the COVID-19 Host Genetics Initiative to investigate similarities and differences in their genetic signals. We identified 9 loci associated with susceptibility (one with two independent GWAS signals; one with an ethnicity-specific signal), 14 associated with severity (one with two independent GWAS signals; two with ethnicity-specific signals) and one harboring two discrepant GWAS signals (one for susceptibility; one for severity). Utilizing colocalization we also identified 45 GTEx tissues that had eQTL(s) for 18 genes strongly associated with GWAS signals in eleven loci (1-4 genes per locus). Some of these genes showed tissue-specific altered expression and others showed altered expression in up to 41 different tissue types. Our study provides insights into the complex molecular mechanisms underlying inherited predispositions to COVID-19-disease phenotypes.

## Introduction

The SARS-CoV-2 virus has infected more than 120 million individuals worldwide between December 2019 and March 2021 and its associated disease (COVID-19) has reportedly caused >2.7 million deaths ^1^. Several risk factors associated with SARS-CoV-2 infection and COVID-19 disease severity have been identified, including older age, sex, ethnicity, blood type, cardiovascular, respiratory and kidney diseases ^2-8^. Most of these risk factors have a well-known genetic component ^9-11^, suggesting that risk of SARS-CoV-2 infection and/or disease severity may also be influenced by the genetic background of an individual. The COVID-19 Host Genetics Initiative (COVID-19 HGI, https://www.covid19hg.org/) is currently leading a public effort worldwide to analyze COVID-19 information for millions of individuals in conjunction with genotype data in order to identify genetic variants associated with SARS-CoV-2 infection as well as COVID-19 hospitalization and disease severity ^12,13^. To date the COVID-19 HGI has conducted a meta-analysis on four phenotypes (very severe respiratory confirmed COVID-19 vs. population, A2; hospitalized vs. non-hospitalized COVID-19 patients, B1; hospitalized vs. population, B2; and COVID-19 patients vs. population, C2) by combining 46 studies worldwide. For each phenotype, four distinct GWASs were performed by either including only individuals from European descent (referred to as European-only hereafter) or individuals from multiple ethnicities (referred to as multi-ethnic hereafter), and using two sets of control individuals either from both the 23andMe and UK Biobank resources or only from 23andMe. Overall, these 16 GWASs identified 15 genomic loci that are significantly associated with SARS-CoV-2 infection and/or COVID-19 severity ^12^, confirming that this disease has a strong underlying genetic component. For several of these loci likely causal variants and their associated target genes have been described and replicated in multiple studies, suggesting that genetic variation contributing to COVID-19 disease is present multiple populations. The HGI dataset, collected from 46 studies worldwide, is ideal to investigate the genetic architecture of COVID-19 disease.

Colocalization is a widely used approach to investigate whether two genetic traits, such as two different GWAS traits mapping into the same locus or expression quantitative trait loci (eQTL) for a specific gene located within a GWAS locus, are associated with the same underlying genetic variants ^14^. This Bayesian method uses p-values of the variants tested for two traits to calculate the posterior probabilities (PP) of four hypotheses at a specific locus: 1) H0: neither trait has a significant association at the tested locus; 2) H1: only the first trait is associated; 3) H2: only the second trait is associated; 4) H3: both traits are associated but the underlying variants are different; and 5) H4: both traits are associated and share the same underlying variants. Therefore, using this approach, it is possible to determine whether the same or different genetic variants within an interval are associated only with SARS-CoV-2 infection, only with COVID-19 disease severity, or both of these related but distinct traits. Likewise, by comparing eQTL and GWAS signals, it is possible to detect genetic variants associated with SARS-CoV-2 infection or COVID-19 disease severity and the differential expression of specific genes in the locus. In this manner, it is possible to exploit large collections of eQTL analyses, which have assessed the associations between genetic variation and gene expression in many human tissues ^15-17^, to identify causal variants in the COVID-19 associated loci and the tissues in which they are functional.

Here, we initially examined the 16 GWASs in the COVID-19 HGI dataset and identified 24 loci that we classified as associated with either SARS-CoV-2 susceptibility or COVID-19 disease severity. We observed that three of these loci likely had an ethnicity-specific component, suggesting that further ethnic-specific studies may identify additional COVID-19-associated loci. Additionally, three loci showed the presence of two independent GWAS signals, indicating that their genetic association with COVID-19 is complex and likely involves multiple independent molecular mechanisms. We next exploited the GTEx collection of eQTL data in 48 human tissues ^15^ to investigate the molecular mechanisms underlying the associations between genetic variation and COVID-19 disease status. We identified variants associated with the altered expression of genes in specific tissues that strongly colocalize with COVID-19-associated variants, thereby providing a potential mechanism underlying the associations between genetic variation, SARS-CoV-2 susceptibility and COVID-19 severity. These findings show that SARS-CoV-2 susceptibility and COVID-19 disease severity have distinct genetic architectures and that ethnicity-specific variation could contribute to observed population differences in these two traits.

## Results

### Genetic architecture of SARS-CoV-2 susceptibility and COVID-19 disease severity

To characterize the similarities and differences of genetic associations between SARS-CoV-2 infection and COVID-19 disease severity phenotypes, we obtained the summary statistics of 16 HGI COVID-19 studies ^12,13^. These 16 GWASs correspond to four distinct phenotypes, one of which is associated with SARS-CoV-2 susceptibility (C2) and three with COVID-19 disease severity (A2, B1, B2). For each phenotype, four GWASs were conducted, of which two were European-only and two were multi-ethnic ^12^. We identified 24 unique loci with genome-wide significant variants (p < 1 × 10^−7^) in at least one study (Table 1, Table S1), of which only one locus on chromosome 3 was significant across all four phenotypes (3:45137109-47151986). To understand if this heterogeneity between phenotypes resulted from different genetic signals or was due to power issues, we applied a colocalization ^14^ approach (Figure 1). By comparing the genetic signal at each locus for each pairwise combination of the 16 GWASs, we were able to determine the posterior probability of whether two studies shared the same underlying genetic associations. GWASs of the same phenotype were more likely to share the same genetic signals (as indicated by high PP-H4 values) than GWASs of different phenotypes (median across all genome-wide significant loci = 0.99 and 0.38, respectively, p = 2.2 × 10^−43^, Mann-Whitney U test, Table S2), indicating that different genomic loci underlie susceptibility to SARS-CoV-2 infection and COVID-19 disease severity phenotypes. We further investigated each of the 24 loci to assign each one as either associated with SARS-CoV-2 susceptibility or COVID-19 severity. We assigned loci associated with susceptibility if two or more of the GWASs in the C2 phenotype (COVID-19 patients vs. population) significantly colocalized between each other (PP-H4 > 0.9). Loci were assigned as associated with disease severity if two or more of the GWASs in either the A2 (Very severe respiratory confirmed COVID-19 vs. population), B1 (Hospitalized vs. non-hospitalized COVID-19 patients) or B2 (Hospitalized COVID-19 patients vs. population) phenotypes significantly colocalized with each other (PP-H4 > 0.9), but none of the GWASs for the C2 phenotype colocalized with one another. Of note, we included all loci for which two or more of the GWASs colocalized with each other in the C2 phenotype as associated with the susceptibility phenotype regardless of whether or not any of the GWASs for the A2 and B2 phenotypes colocalized with one another, as the use of population-based controls in these latter two phenotypes resulted in the capture of both severity and susceptibility. Conversely, B1 primarily captured disease severity, as its controls included only non-hospitalized COVID-19 patients. Across all populations (i.e., the two European-only and the two multi-ethnic GWASs) we identified seven loci associated with susceptibility (Figure 1, first row, Table S2) and 11 loci associated with disease severity (but not susceptibility) (Figure 1, second and third row). Three loci, including two associated with susceptibility (C2) and one with disease severity (B2), showed heterogeneity in strength of colocalization between the European-only and multi-ethnic GWASs for one or more phenotypes (Figure 1, fourth row, Figure 2A-I), indicating the presence of ethnicity-specific signals. Three loci had both high PP-H3 and PP-H4 scores from the colocalization analysis, indicating the presence of two independent GWAS signals (i.e., independent causal variants, Figure 1, fifth row, Figure 2J-R). These analyses show that different intervals in the genome are associated with SARS-CoV-2 susceptibility and COVID-19 disease severity, and that the genetic associations within these intervals can be ethnicity-specific and/or complex involving multiple independent GWAS signals.

**Table 1:**
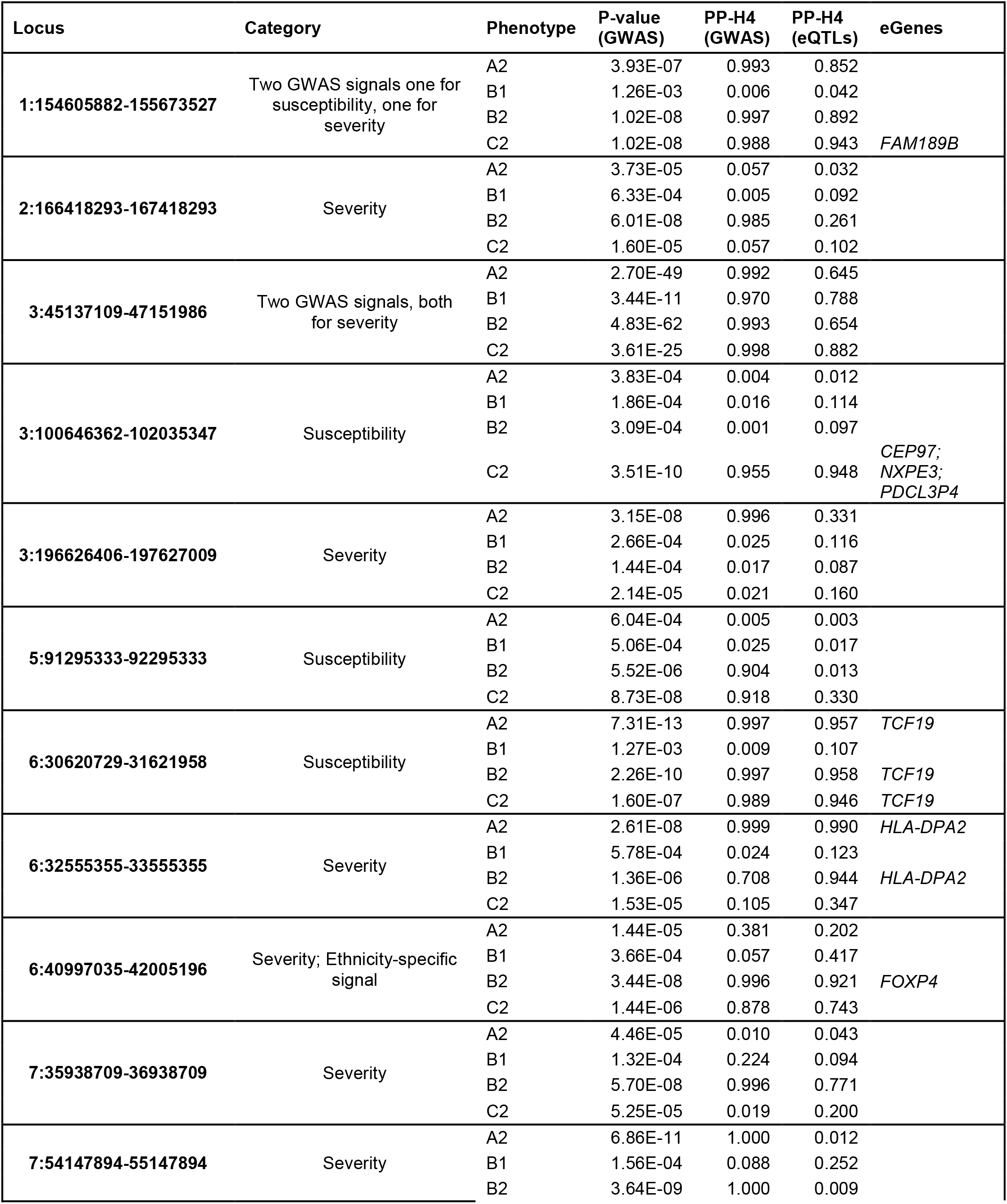
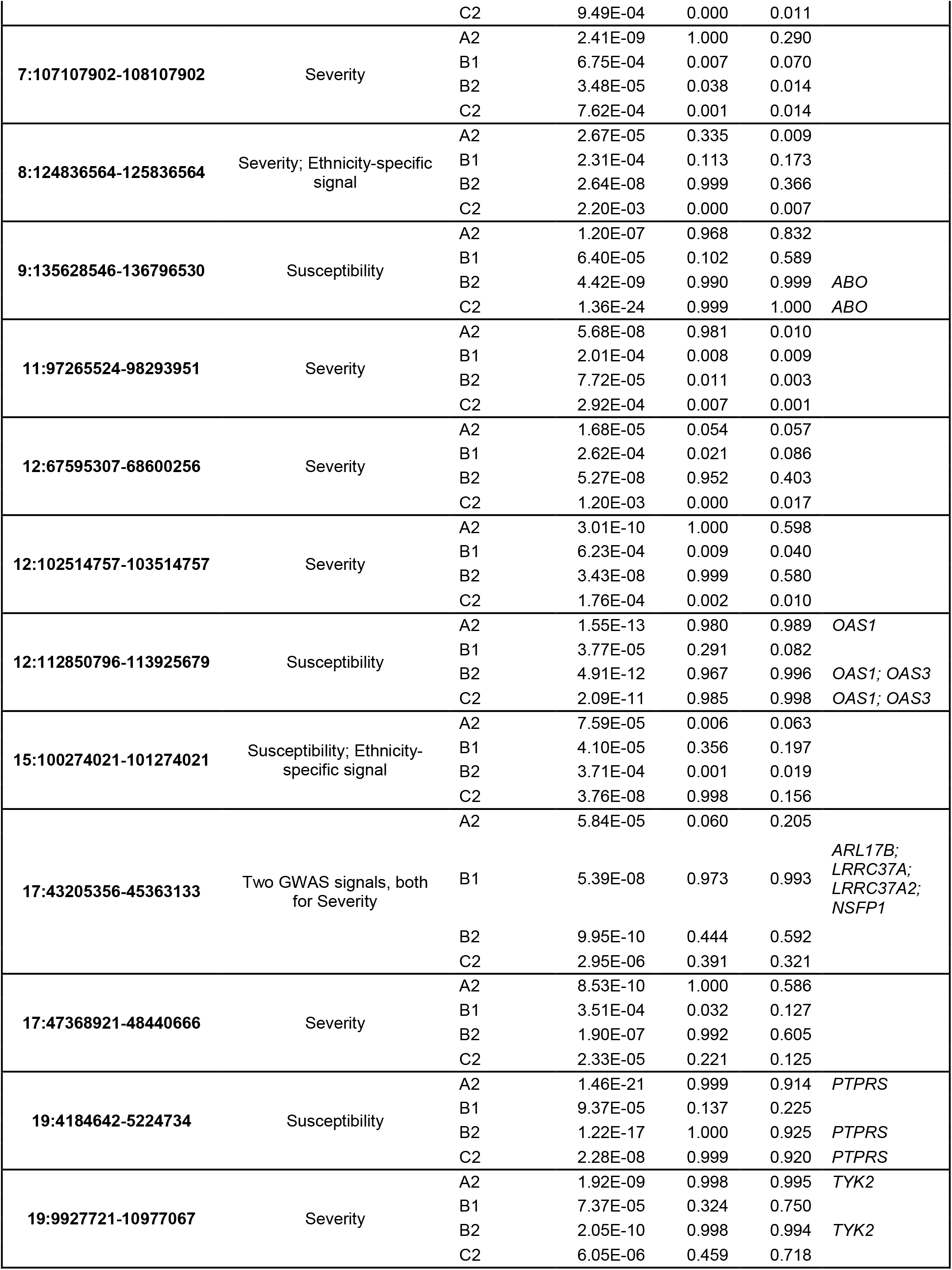
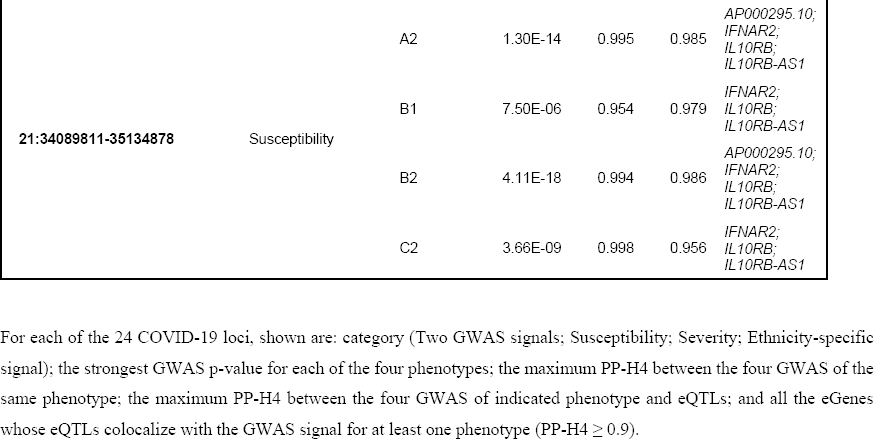
Genome-wide significant loci.

**Figure 1:**
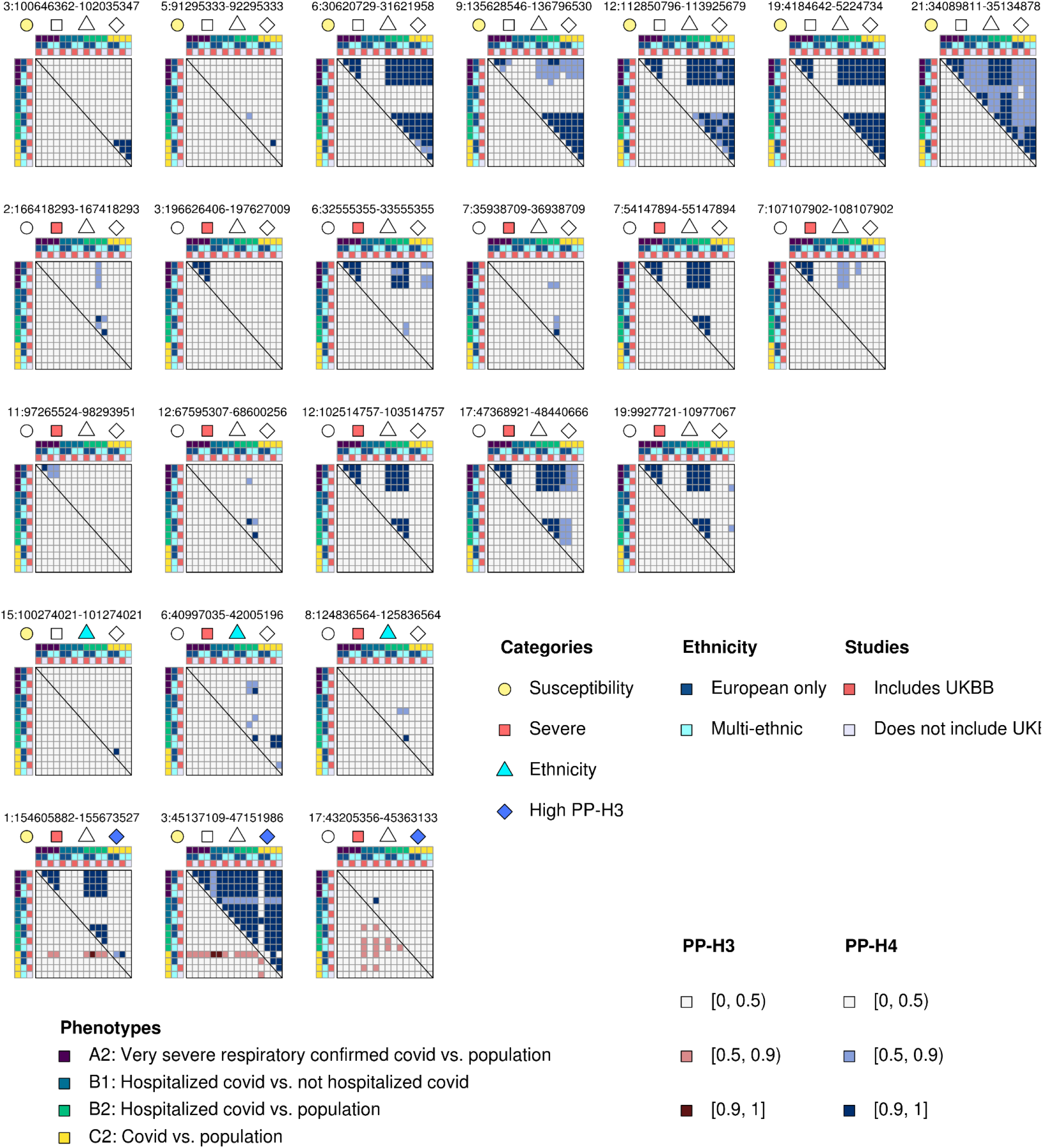
Colocalization of GWAS signals across four COVID-19 phenotypes. Heatmaps showing the posterior probability of association between each pairwise combination of the 16 GWAS studies at each of the 24 genome-wide significant loci across the four phenotypes (A2, B1, B2, C2). In each heatmap, studies are sorted by phenotype (colors are indicated on the first row and first column), ethnicity (second row and second column: dark blue is for European only, cyan is for multi-ethnic) and type of controls (third row and third column: red indicates studies that use both 23andMe and UK Biobank (UKBB) individuals as controls, while gray indicates studies that do not use UK Biobank individuals). Each heatmap is divided into two triangles: the upper right shows PP-H4 (blue scale), whereas the bottom left shows PP-H3 (red scale). The symbols below the coordinates of each locus indicate whether each locus is associated with susceptibility (yellow circle), severity (red square), high PP-H3 (blue diamond) or ethnicity-specific (cyan triangle). Top row are seven loci associated only with infection. Second and third rows are 11 loci associated only with severity. Third row are three loci containing ethnicity-specific signals. Fourth row are three loci with two independent GWAS signals (high PP-H3 between studies).

**Figure 2:**
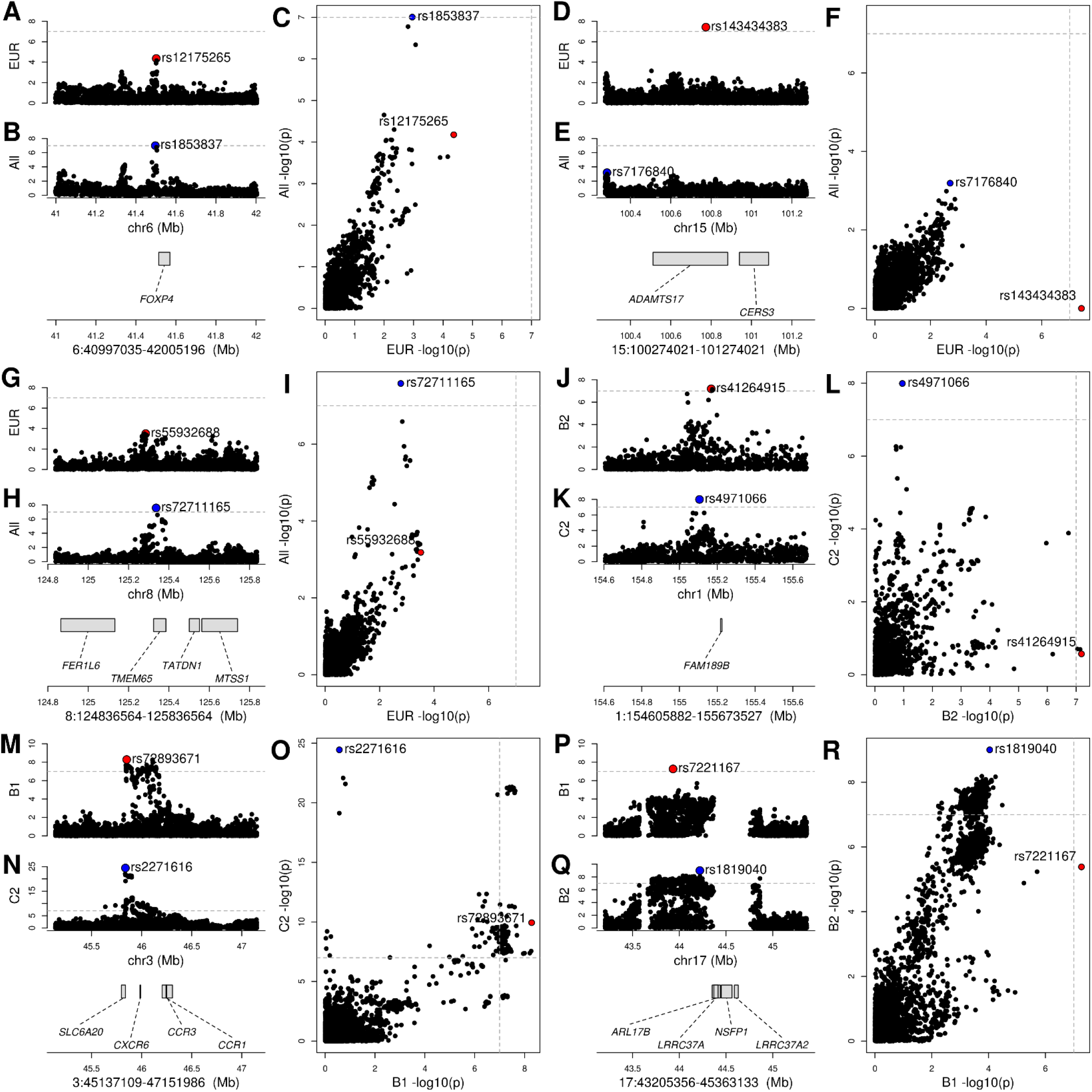
Colocalization between COVID-19 phenotypes identifies multiple GWAS signals in three loci. (A, B, C) GWAS signal at the 6:40997035-42005196 locus for (A) individuals of European descent (*COVID19_HGI_B2_ALL_eur_leave_ukbb_23andme*) and (B) all individuals from multiple ethnicities (*COVID19_HGI_B2_ALL_leave_UKBB_23andme*). Only when testing all individuals from multiple ethnicities, p-values in this locus are genome-wide significant (Table 1), suggesting that this locus is associated with individuals of non-European descent. Indeed, the lead variant at this locus (rs1853837) is less common in Europeans than East Asians, Admixed Americans, Africans or Ashkenazi Jewish. (C) The scatterplot showing the –log10 (p-value) calculated in these two GWAS for all variants in the 6:40997035-42005196 locus confirms that the GWAS signal is absent when testing only individuals of European descent. (D, E, F) GWAS signal at the 15:100274021-101274021 locus for (D) individuals of European descent (*COVID19_HGI_C2_ALL_eur_leave_23andme*) and (E) all individuals from multiple ethnicities (*COVID19_HGI_C2_ALL _leave_23andme*). The lead variant in individuals of European descent (rs143434383) is rare (1.1% allele frequency) and was not tested in other ethnicities (allele frequency < 1%), indicating that the signal at this locus is likely European-specific. (F) The scatterplot showing the –log10 (p-value) calculated in these two GWAS for all variants in the 15:100274021-101274021 locus confirms that the GWAS signal is present when testing only individuals of European descent. (G, H, I) GWAS signal at the 8:124836564-125836564 locus for (G) individuals of European descent (*COVID19_HGI_B2_ALL_eur_leave_23andme*) and (H) all individuals from multiple ethnicities (*COVID19_HGI_B2_ALL _leave_23andme*). Only when testing all individuals from multiple ethnicities, p-values in this locus are genome-wide significant, suggesting that this locus is associated with individuals of non-European descent. Indeed, the lead variant at this locus (rs72711165) is less common in Europeans than East Asians or Africans. (I) The scatterplot showing the –log10 (p-value) calculated in these two GWAS for all variants in the 8:124836564-125836564 locus confirms that the GWAS signal is absent when testing only individuals of European descent. (J, K, L) GWAS signal at the 1:154605882-155673527 locus for (J) B2 phenotype (hospitalized COVID-19 patients vs. population: *COVID19_HGI_B2_ALL_eur_leave_23andme*) and (K) C2 phenotype (COVID-19 patients vs. population: *COVID19_HGI_C2_ALL_eur_leave_23andme*). The two GWAS have different lead variants (rs41264915 and rs4971066, 64.5 kb apart, not in LD) and GWAS signals. (L) The scatterplot showing the –log10 (p-value) calculated in these two studies for all variants in the 1:154605882-155673527 locus confirms that different genetic variants have significant p-values, suggesting the presence of two independent signals. (M, N, O) GWAS signal at the 3:45137109-47151986 locus for (M) B1 phenotype (hospitalized COVID-19 patients vs. non-hospitalized COVID-19 patients: *COVID19_HGI_B1_ALL_eur_leave_23andme*) and (N) C2 phenotype (COVID-19 patients vs. population: *COVID19_HGI_C2_ALL_eur_leave_23andme*). The two studies have different lead variants (rs72893671 and rs2271616, ∼12 kb apart, not in LD) and GWAS signals. (O) The scatterplot showing the –log10 (p-value) calculated in these two GWAS for all variants in the 3:45137109-47151986 locus confirms that different genetic variants have significant p-values, suggesting the presence of two independent signals. (P, Q, R) GWAS signal at the 17:43205356-45363133 locus for (P) B1 phenotype (hospitalized COVID-19 patients vs. non-hospitalized COVID-19 patients: *COVID19_HGI_B1_ALL_leave_23andme_20210107*) and (Q) B2 phenotype (Hospitalized COVID-19 patients vs. population: *COVID19_HGI_B2_ALL_leave_23andme_20210107*). The two GWAS have different lead variants (rs7221167 and rs1819040, ∼300 kb apart, not in LD) and GWAS signals. (R) The scatterplot showing the –log10 (p-value) calculated in these two studies for all variants in the 17:43205356-45363133 locus confirms that different genetic variants have significant p-values, suggesting the presence of two independent signals.

### Colocalization with eQTLs

To identify genes whose differential expression is associated with SARS-CoV2 infection and COVID-19 severity, we obtained eQTL data in 48 GTEx tissues ^15^ for 679 genes that map to the 24 GWAS loci and performed a colocalization analysis ^14^ between each eQTL and GWAS signal. Eleven of the 24 loci had GWAS signals in one or more of the four phenotypes that colocalized (PP-H4 > 0.9) with eQTLs for 20 genes (Figure 3, Figure 4, Figure 5, Table S3). The GWAS signals in seven of these loci colocalized with eQTLs for a single gene, of which four eQTLs were present in a single tissue and three in four or more tissues (range: 4-21). In the other four loci the GWAS signals colocalized (PP-H4 > 0.9) with eQTLs for 13 genes (range: 2-4 per loci), with each eQTL showing colocalization in one to 41 tissues. Further examination of these four loci showed that in two (3:100646362-102035347 and 21:34089811-35134878) there was a single eQTL associated with the differential expression of multiple genes across different tissues (Figure 4A,B), whereas the other two (12:112850796-113925679 and 17:43205356-45363133) showed high PP-H3, suggesting the presence of two distinct eQTL signals (Figure 4C,D).

**Figure 3:**
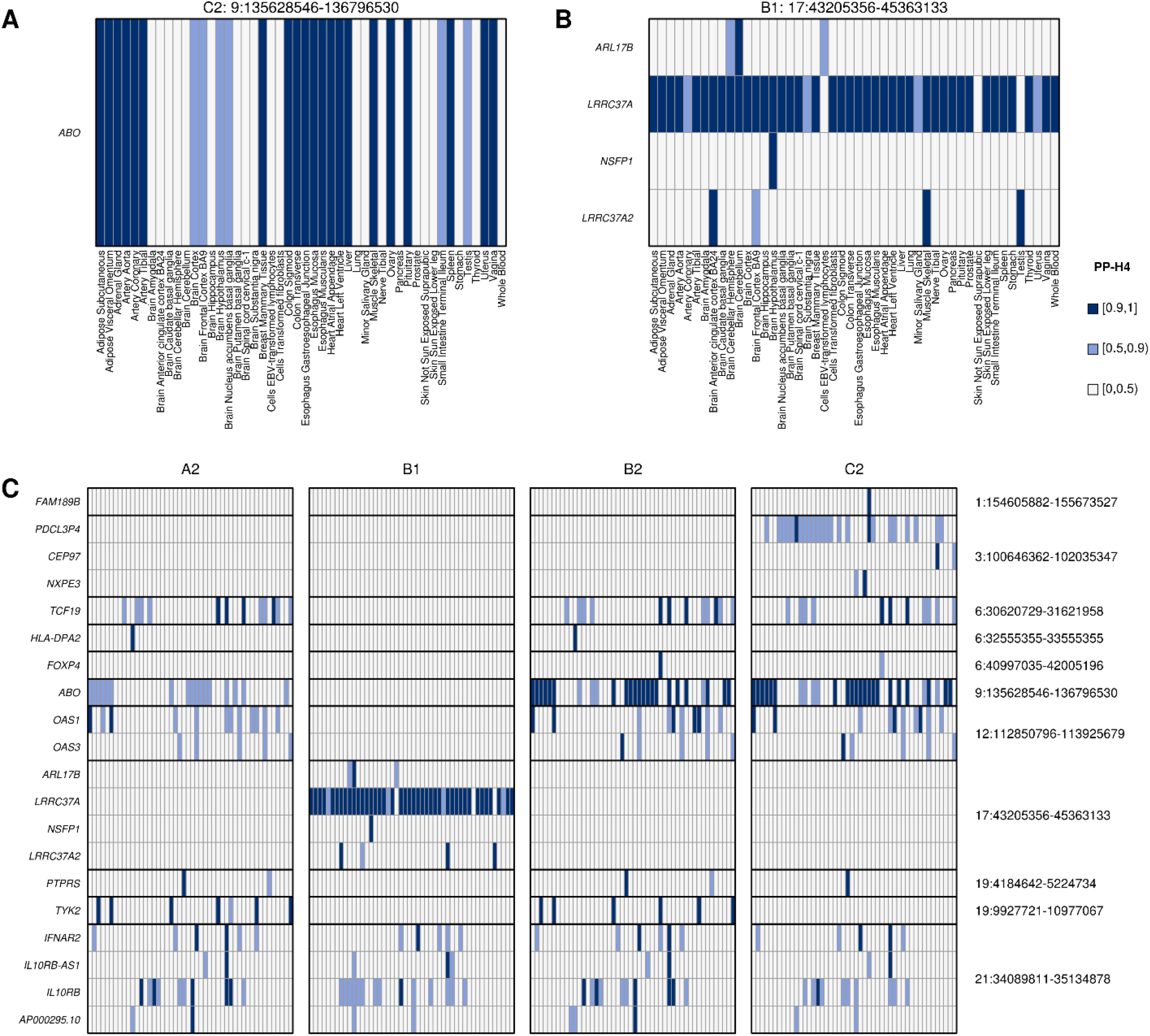
Colocalization of COVID-19 GWAS signals and eQTLs in 48 tissues. (A,B) Heatmaps showing the posterior probability of eQTLs in indicated tissue with (A) the 9:135628546-136796530 locus C2 phenotype (COVID-19 patients vs. population) and (B) the 17:43205356-45363133 locus B1 phenotype (Hospitalized vs. non-hospitalized COVID-19 patients). Gene(s) with eQTLs in the indicated tissue that colocalize with the GWAS signal in the indicated interval with PP-H4 ≥ 0.9 trait are shown in dark blue; or in light blue if 0.5 ≤ PP-H4 < 0.9. (C) Heatmap showing the posterior probability of eQTLs colocalize with GWAS signals for each of the four COVID-19 phenotypes: 1) A2 = Very severe respiratory confirmed COVID-19 vs. population; 2) B1 = Hospitalized vs. non-hospitalized COVID-19 patients; 3) B2 = Hospitalized COVID-19 patients vs. population; and 4) C2 = COVID-19 patients vs. population. Each column represents a tissue, sorted in alphabetical order as shown in (A,B). Each cell in the heatmaps show the highest PP-H4 calculated across the four GWAS (Figure 1) for the indicated phenotype.

**Figure 4:**
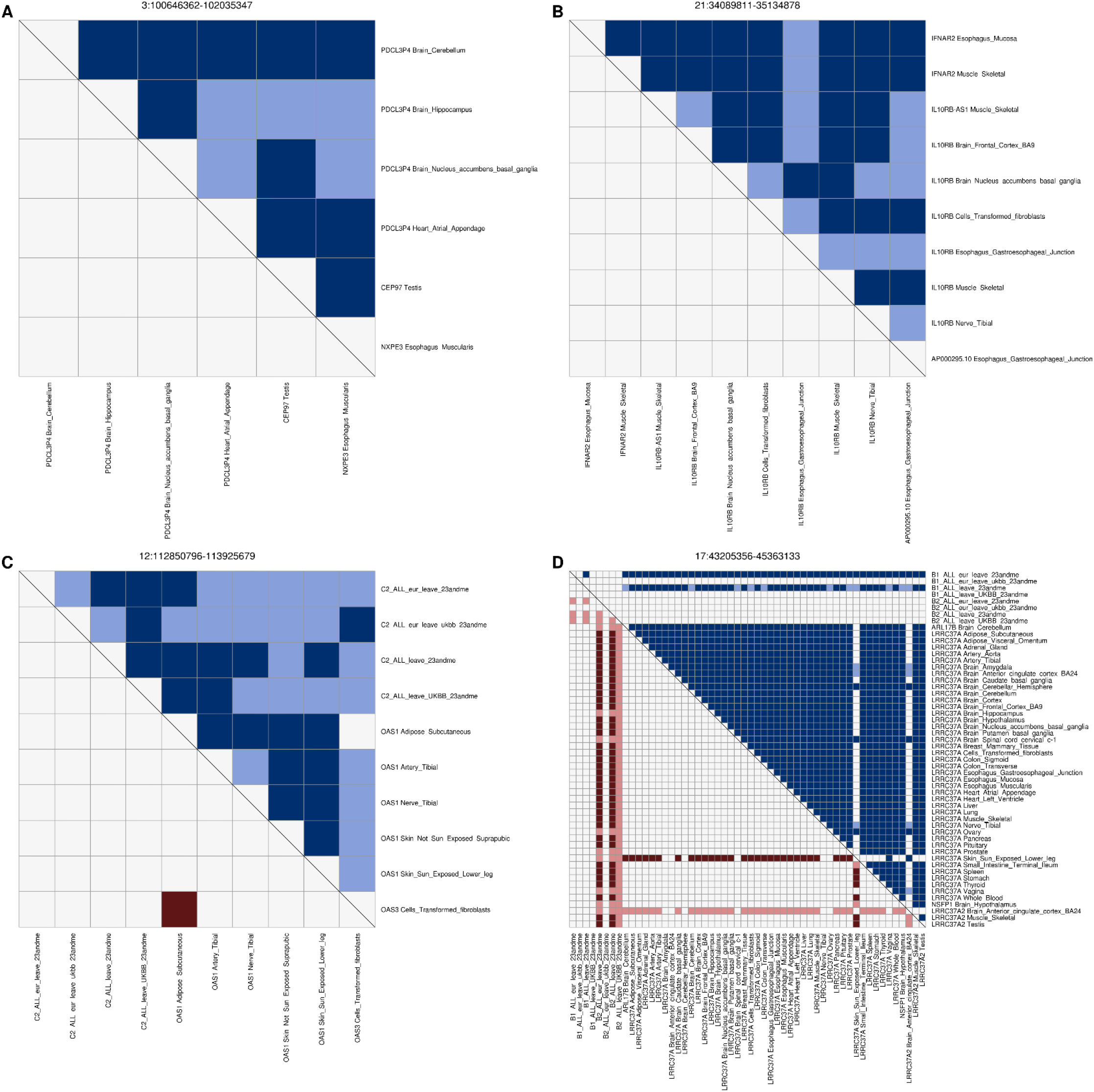
eQTL signals for multiple genes at the same GWAS locus. Heatmaps showing the colocalization of tissue eQTLs for multiple genes within the same COVID-19 GWAS loci. In panels (A) and (B) the same eQTL is associated with the differential expression across tissue types of the respective three and four genes in the locus. In panels (C) and (D) colocalization between eQTLs and GWAS are shown: (C) C2 phenotype; and (D) B1 and B2 phenotypes. In each heatmap, the upper triangle represents PP-H4 values, where dark blue indicates PP-H4 ≥ 0.9 and light blue indicates 0.5 ≤ PP-H4 < 0.9; the lower triangle represents PP-H3 values, where dark red indicates PP-H3 ≥ 0.9 and light blue indicates 0.5 ≤ PP-H3 < 0.9.

**Figure 5:**
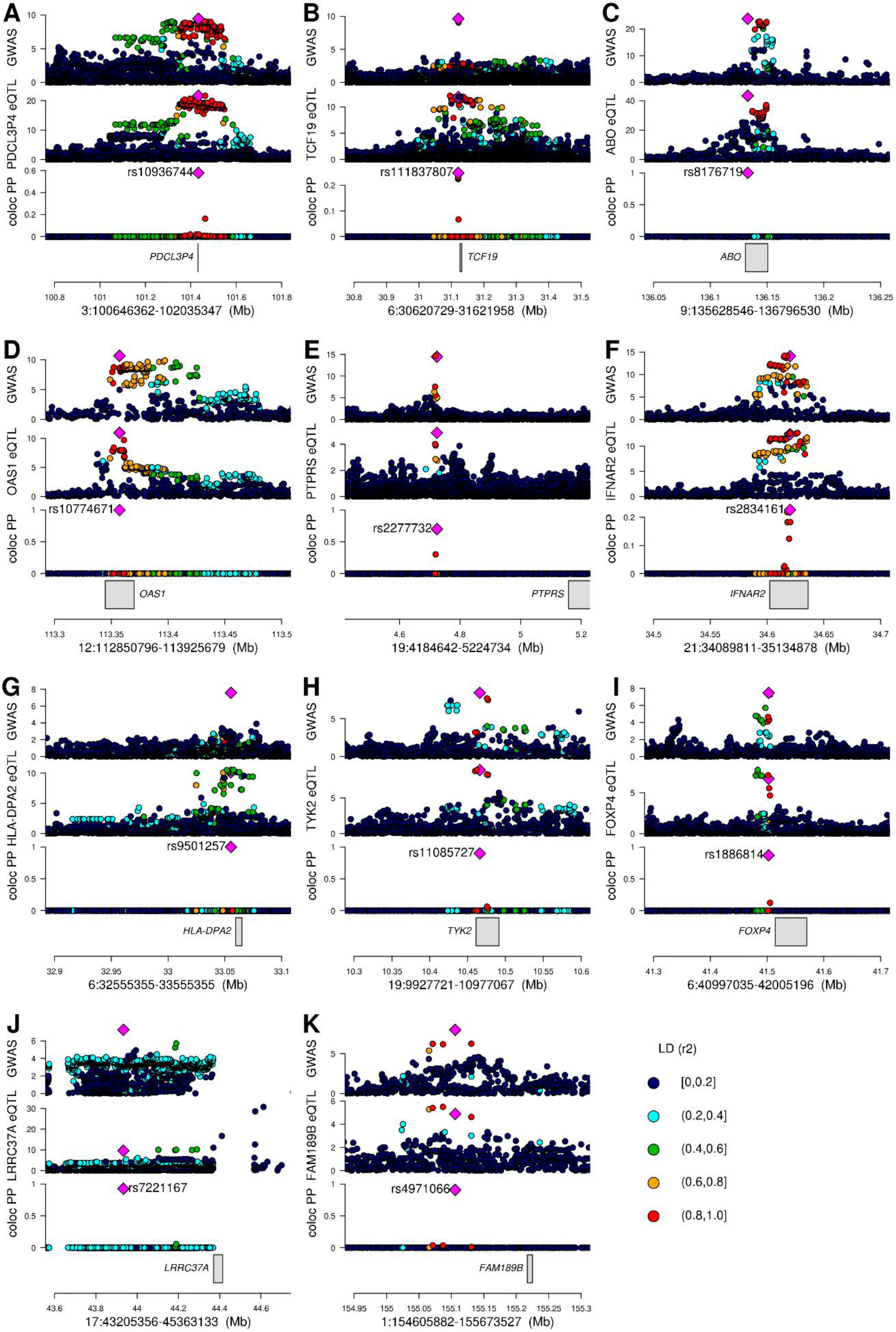
Colocalizations between COVID-19 GWAS and tissue eQTL signals. Association between GWAS and eQTL signals in 11 loci: (A) locus 3:100646362-102035347, phenotype C2 GWAS and *PDCL3P4* eQTL in Brain Cerebellum; (B) locus 6:30620729-31621958, phenotype B2 GWAS and *TCF19* eQTL in Lung; (C) locus 9:135628546-136796530, phenotype C2 GWAS and *ABO* eQTL in Adipose Visceral Omentum; (D) locus 12:112850796-113925679, phenotype C2 GWAS and *OAS1* eQTL in Adipose Subcutaneous; (E) locus 19:4184642-5224734, phenotype B2 GWAS and *PTPRS* eQTL in Colon Sigmoid; (F) locus 21:34089811-35134878, phenotype B2 GWAS and *IFNAR2* eQTL in Muscle Skeletal; (G) locus 6:32555355-33555355, phenotype A2 GWAS and *HLA-DPA2* eQTL in Brain Cerebellum; (H) locus 19:9927721-10977067, phenotype A2 GWAS and *TYK2* eQTL in Adrenal Gland; (I) locus 6:40997035-42005196, phenotype B2 GWAS and *FOXP4* eQTL in Lung; (J) locus 17:43205356-45363133, phenotype B1 GWAS and *LRRC37A* eQTL in Pancreas; (K) locus 1:154605882-155673527, phenotype C2 GWAS and *FAM189B* eQTL in Heart Atrial Appendage. For each locus, four panels are presented: 1) GWAS p-values for indicated COVID-19 phenotype; 2) eQTL p-values; 3) posterior probability of colocalization of each variant, calculated using coloc, color coded based on linkage disequilibrium in individuals of European descent (r^2^) calculated using LDlinkR ^57^; and 4) gene loci. Since the *FOXP4* locus was associated with non-European individuals, LD in panel (I) is shown for East Asians.

Of the 48 tissues examined for colocalization between eQTLs and COVID-19 GWAS signals, 45 had eQTLs that colocalized with at least one GWAS signal, of which 18 colocalized with two or more (mean: 2.1; median: 2; range: 0 to 7). These 45 tissues include several that are associated with COVID-19 symptoms, such as lung (associated with cough and shortness of breath: *FOXP4, LRRC37A, TCF19* and *TYK2*), whole blood (immune response: *LRRC37A* and *TYK2*), skeletal muscle (muscle fatigue and body aches: *ABO, IFNAR2, IL10RB, IL10RB-AS1, LRRC37A, LRRC37A2* and *TCF19*), digestive system (nausea and diarrhea: *ABO, LRRC37A, PTPRS, AP000295*.*10, IL10RB, IFNAR2* and *NXPE3*) and brain cortex (loss of smell and taste, cognitive defects: *LRRC37A* and *LRRC37A2*, Table S3). Surprisingly, most of the GWAS signals did not colocalize with eQTLs in whole blood or lung, the two tissues that have been described as the most affected by COVID-19. These findings show that causal variants underlying COVID-19 GWAS signals colocalize with regulatory variants associated with the differential expression of gene(s), of which some show tissue-specific altered expression and others show altered expression across many different tissue types.

### Loci containing associations between SARS-CoV2 susceptibility and eQTLs

We found seven loci that showed associations with the C2 phenotype (SARS-CoV2 susceptibility), six of which colocalized with eQTLs.

The GWAS signal for interval 3:100646362-102035347 colocalized with an eQTL signal of three genes in four distinct tissues (*PDCL3P4*: cerebellum, hippocampus, nucleus accumbens basal ganglia and atrial appendage; *CEP97*: testis; and *NXPE3*: esophagus muscularis, Figure 3C, Figure 4A, Figure 5A), suggesting the causal variant in this locus affects SARS-CoV-2 susceptibility by altering the expression of one or more of these genes in a tissue-specific manner.

In the 6:30620729-31621958 locus the GWAS signal colocalized with *TCF19* eQTLs in lung, skeletal muscle, pituitary gland and testis (Figure 3C, Figure 5B). This gene was previously described as functionally associated with COVID-19 ^18,19^, and is involved in lung cancer ^20^. Given the presence of multiple variants in high LD, we were not able to identify one single likely causal variant, but observed four with posterior probability of being causal > 0.2.

eQTLs for the *ABO* gene in 21 tissues colocalized with COVID-19 susceptibility (Figure 3A,C). This gene is expressed in erythrocytes, endothelial and epithelial cells ^21,22^ and polymorphisms at two variants in its sequence (rs8176746 and rs8176719) confer the O, A, B and AB blood types ^23^. The variant with highest PP of being causal was rs8176719 across all 21 tissues (Figure 5C), which distinguishes the O blood type (reference allele) from the A and B blood types (C insertion), which is consistent with the fact that blood type O is a factor of reduced risk for SARS-CoV-2 infection ^24^.

The 12:112850796-113925679 locus encodes a cluster of three 2’-5’-Oligoadenylate Synthetases (*OAS1, OAS2* and *OAS3*) and harbors eQTLs for *OAS1* in five tissues (Figure 5D) and for *OAS3* in transformed fibroblasts. The five *OAS1* eQTLs (one per tissue) colocalized with each other, suggesting the presence of one single causal variant affecting the expression of OAS1, and also colocalized with the C2 COVID-19 GWAS signal (Figure 4C). While the *OAS3* eQTL was stronger than the eQTL for *OAS1* (p < 10^−47^ for *OAS3*, p > 10^−12^ for *OAS1*, Table S3) it did not colocalize with either the *OAS1* eQTLs or the C2 COVID-19 signal. These data suggest that, while all three OAS genes play a critical role in cellular innate antiviral response ^25^, differential expression of *OAS1* is likely causally implicated in COVID-19 susceptibility. These findings are in agreement with previous studies showing that high plasma OAS1 levels are associated with reduced COVID-19 susceptibility and severity ^26^.

In the 19:4184642-5224734 locus the GWAS signal colocalized with an eQTL in sigmoid colon for *PTPRS* (Figure 3C, Figure 5E). The eQTL is 481 kb downstream of *PTPRS* and hence likely in a distal regulatory element. Interestingly, its protein product was shown to interact with the membrane (M) protein of SARS-CoV-2 ^27,28^, which likely glycosylates the spike protein ^29^.

The 21:34089811-35134878 locus colocalized with eQTLs for four genes (*IFNAR2, IL10RB, IL10RB-AS1* and *AP000295*.*10*) in nine tissues (skeletal muscle, esophagus mucosa, gastroesophageal junction, frontal cortex, nucleus accumbens basal ganglia, tibial nerve and transformed fibroblasts, Figure 4B). Interestingly, most of these eQTLs colocalized with each other (Figure 5F), suggesting that a single causal variant underlies the altered expression of all four genes in the different tissue types and COVID-19 susceptibility. We were not able to identify one single likely causal variant due to the LD structure, but observed five variants with posterior probability of being causal between 0.1 and 0.3.

### Associations between COVID-19 severity and eQTLs

We found 11 loci that showed associations with COVID-19 severity, including two with colocalizing eQTLs.

The 6:32555355-33555355 locus overlapped with the major histocompatibility complex (MHC) and the GWAS signal colocalized with an eQTL in the cerebellum for *HLA-DPA2* (Figure 5G), a pseudogene member of the MHC class II ^30,31^. The most likely causal variant (rs9501257) underlying both of these genetic signals is located in the 3’ UTR of *HLA-DPB1*.

In the 19:9927721-10977067 locus, the GWAS signal for the COVID-19 A2 and B2 phenotypes colocalized with eQTLs for *TYK2* in seven tissues, including adrenal gland, tibial artery, breast, lung, tibial nerve, skin and whole blood (Figure 3C, Figure 5H). The most likely causal variant (rs11085727) was the lead variant in a GWAS signal for neutrophil percentage, systemic lupus erythematosus and thyroid medication ^32-34^, suggesting the involvement of this locus in inflammation and the cytokine storm associated with the most severe COVID-19 cases ^35,36^.

### COVID-19 response has ethnicity-specific components

We observed three loci with ethnicity-specific associations, including one that colocalized with eQTLs.

The 6:40997035-42005196 locus showed colocalization between multi-ethnic GWAS, but not between European-only GWAS (Figure 1, Figure 2A,B,C). Interestingly, the lead GWAS variant in this locus (rs1886814) is less common in Europeans (gnomAD allele frequency = 2.4%) than in East Asians (37.3%), Admixed Americans (18.9%), or South Asians (11.8%) ^37^ and was found to be associated with COVID-19 in East Asians by the COVID-19 HGI ^12,13^. The lung-specific eQTL for *FOXP4* (rs1886814) colocalized with the COVID-19 B2 phenotype (Figure 5I) and is in LD with an emphysema-associated variant (rs2894439, r^2^ = 0.7) ^38,39^, suggesting a potential role for this gene in disease severity.

A second locus (8:124836564-125836564) containing an ethnicity-specific signal based on colocalization has also been reported by the COVID-19 HGI as obtaining genome-wide significance only in East Asian populations with the leading variant (rs72711165) at lower frequency in other populations ^12,13^.

The lead variant in the third locus 15:100274021-101274021 is rare in individuals of European descent (rs143434383, 1.1% allele frequency) and was not tested in the multi-ethnic studies because it is even rarer in other ethnicities (allele frequency < 1%). Although our results suggest that this is a European-only GWAS signal because of the allele frequency of this variant further studies are required to confirm this observation.

### Independent GWAS signals at three loci

We identified three COVID-19-associated loci that each showed two independent GWAS signals.

In the 1:154605882-155673527 locus, two C2 GWASs showed high PP-H4 with each other but PP-H3 of varying strength with other GWASs (Figure 3C) indicating the presence of two independent GWAS signals, one for susceptibility and one for severity. The GWAS signal associated with COVID-19 susceptibility in this interval colocalized with an eQTL for *FAM189B* in cardiac tissues (atrial appendage, Figure 5K). Although this gene’s function has not been well-characterized, it has been shown to be associated with triglyceride levels in the blood ^40^. Furthermore, recent evidence showed that the FAM189B protein product can bind to the SARS-CoV-2 envelope (E) protein ^27,28^, which contributes to virus assembly and is important for the virus pathogenicity ^41^. These observations, combined with the role of triglyceride levels as prognostic markers for COVID-19 ^42-45^, suggest that *FAM189B* function is likely involved in the association between lipid levels and COVID-19.

Most of the GWASs in the 3:45137109-47151986 locus showed high PP-H4 with each other indicating a signal for susceptibility across all populations. However, one C2 GWAS showed strong PP-H3 signals with two B1 GWASs and varying strength PP-H3 signals with other GWASs (Figure 3C) indicating the presence of a second independent GWAS signal most likely also for susceptibility. Neither GWAS signal in this interval colocalized with any eQTLs.

One locus (17:43205356-45363133) showed different GWAS signals between the B1 and B2 phenotypes (PP-H3 = 0.81, Figure 1, Table S2), suggesting the presence of two severity-associated signals. While we observed two potential GWAS signals for two phenotypes (B1, B2), we found that eQTLs colocalized only with the B1 phenotype (Figure 3B,C, Figure 4D). The B1 GWAS signal colocalized with *LRRC37A* eQTLs in 41 tissues, *ARL17B* in cerebellum, *NSFP1* in hypothalamus and *LRRC37A2* in anterior cingulate cortex, skeletal muscle and testis (Figure 5J, Table S3). Colocalization between eQTL signals (Figure 4D) showed the presence of a second eQTL for *LRRC37A* in skin and *LRRC37A2* in anterior cingulate cortex, which had moderate to high PP-H3 with all the other eQTLs and high PP-H4 colocalization only with one European-only B1 GWAS. While these observations indicate that this association might have an ethnicity-specific component, further studies will be required to characterize the extent to which this locus is associated with different populations. While the role the genes in this interval play in COVID-19 disease are unknown, the two variants with highest posterior probability of being causal for both the *LRRC37A* eQTLs and COVID-19 hospitalization (rs7221167 and rs7225002, Table S3) were strongly associated with Parkinson’s disease (p = 1 × 10^−37^ and 2 × 10^−40^, respectively) ^46^, suggesting a complex association between this locus and the neurological effects of COVID-19.

## Discussion

The COVID-19 pandemic has united the focus of the broad biomedical scientific community to improve our understanding of the mechanisms underlying disease susceptibility and severity. The COVID-19 HGI has collected genetic and COVID-19 status information from ∼2 million individuals across 46 studies ^12,13^, making COVID-19 one of the best-powered GWAS studies ever conducted. Standard approaches to characterize GWAS summary statistics focus on obtaining all the genomic loci where variants have p-value < 5 × 10^−8^ and identifying the most likely causal gene by proximity to the lead variant. Here, we used a colocalization method to conduct pairwise comparisons of 16 GWAS studies for four COVID-19 disease phenotypes to investigate the similarities and differences in their genetic signals. This resulted in four possible annotations of each locus. 1) We annotated a locus as “COVID-19 susceptibility” if it was significant (p-value < 10^−7^) in at least one of the four C2 GWASs and if at least two C2 studies colocalized. These loci may be significant and colocalize with severity phenotypes. However, since severity phenotypes A2 and B2 used the same general population as control, these signals likely reflect susceptibility rather than severity. 2) All loci for which at least two GWASs in one of the three severity phenotypes (A2, B1 or B2) colocalized, but not with susceptibility, were annotated as “COVID-19 severity. 3) In certain cases, not all the studies from the same phenotype were significant and colocalized. The loci where the European-only GWAS did not colocalize with the multi-ethnic only GWAS from the same phenotype, were labeled as “ethnicity-associated”. 4) Three studies with high PP-H3 at one locus indicated the presence of two distinct GWAS signals. These loci were labeled as having “two GWAS signals”. Using these annotations, we were able to confirm the findings by the COVID-19 HGI at multiple loci and to discover novel associations at previously uncharacterized loci. Overall, these analyses show that the associations between GWAS signals across the four COVID-19 phenotypes are heterogeneous indicating that different genetic variants are likely associated with SARS-CoV-2 susceptibility and COVID-19 severity. We also applied the same colocalization method to compare the COVID-19 GWASs and eQTLs signals, based on the assumption that genetic variants may be associated with disease because they overlap elements that regulate the expression of specific genes. GTEx provided us with the unique opportunity to investigate the associations between genotype, gene expression and COVID-19 in 48 tissues, rather than only lung and blood eQTLs. This resulted in the detection of eQTLs associated with COVID-19 in 45 tissues, providing a putative explanation of the systemic effects of SARS-CoV-2 infection.

One of the best-characterized COVID-19 susceptibility loci is *ABO*. We confirmed that its eQTLs in 27 tissues colocalized with COVID-19 susceptibility and that the most likely causal variant (posterior probability > 0.99) for both eQTLs and GWASs was the non-synonymous variant rs8176719, which distinguishes the O blood type from the A and B blood types, as multiple studies previously described ^12,18^. Likewise, we confirmed lung-specific eQTLs for *FOXP4* to colocalize with COVID-19 severity. This locus, as well as the 8:124836564-125836564 locus, had been described as East Asian-specific by the COVID-19 HGI ^12^. Although we were not able to obtain summary statistics for East Asian populations, our results show that these loci colocalize only between multi-ethnic studies, but not with European-only, suggesting that our approach allows for the detection of ethnicity-specific associations. For many loci we identified the same associated genes as described by the COVID-19 HGI study. For example, we found that one *OAS1* eQTL in multiple tissues (rs10774671), which affects its splice site, is likely causal for COVID-19. However, while the COVID-19 HGI study found this interval to be associated with severity, our analysis classified this locus as associated with susceptibility, which is consistent with the role of OAS1 in preventing infection by activating RNase L, which promotes viral RNA degradation ^47^. Furthermore, in the 21:34089811-35134878 locus we found eQTLs for *IL10RB, IL10RB-AS1* and *AP000295*.*10*, in addition to *IFNAR2*, which colocalized with COVID-19. While both *IFNAR2* and *IL10RB* have been described as likely associated with COVID-19, our results show that the genotypes of several variants in high LD are associated with the expression of these four genes (two protein coding: *IFNAR2* and *IL10RB*; one antisense: *IL10RB-AS1*; and one long noncoding RNA of unknown function: *AP000295*.*10*) and, in turn, with COVID-19 susceptibility, suggesting that all four genes may be involved in the disease.

For three loci we identified two independent GWAS signals associated with COVID-19. eQTLs for *FAM189B*, a gene whose protein product is involved in lipid metabolism and binds the SARS-CoV-2 envelope (E) protein ^27,28^, colocalized with one of two GWAS signals at the 1:154605882-155673527 locus. Given the role of triglyceride levels as a prognostic marker for COVID-19 ^42-45^, *FAM189B* function is likely involved in the association between lipid levels and COVID-19. The 17:43205356-45363133 locus was the most complex containing two distinct GWAS signals and two distinct eQTL signals. The B1 GWAS and eQTL signal for *LRRC37A* strongly colocalized across 41 tissues. Of note, this locus contains an inversion polymorphism, which may exist as a direct or inverted (a 1.5 Mb inversion) haplotype ^48,49^, and has been associated with multiple diseases, including Parkinson’s disease ^46^, Alzheimer’s disease ^52^, Koolen-De Vries Syndrome, which is associated with hypotonia, seizures and cardiac defects ^50^, and progressive supranuclear palsy, which is a neurodegenerative disease that affects movement, cognitive ability, speech and vision ^51^. Interestingly, the two variants with the highest posterior probability of being causal for both the B1 phenotype and eQTLs (rs7221167 and rs7225002, Table S3) were both strongly associated with Parkinson’s disease (p = 1 × 10^−37^ and 2 × 10^−40^, respectively) ^46^. While further studies are required, the variants associated with neurological disorders, the expression of four genes in many tissues and the association with COVID-19 hospitalization suggest that this chromosome 17 locus may be involved in the medium- and long-term neurological effects of SARS-CoV-2 infection ^53-55^.

In summary, our study describes the associations between COVID-19 disease phenotypes, genetic variation and gene expression in 48 human tissues, thereby providing insights into the molecular mechanisms underlying inherited predispositions to SARS-CoV-2 infection and COVID-19 severity. While we described several genes with strong associations with COVID-disease phenotypes, further studies are needed to understand the functional mechanisms underlying these associations, as in several cases the genes do not have well-known functions.

## Methods

### GWAS summary statistics processing

We downloaded the summary statistics on GRCh37 coordinates of 16 GWAS studies from the COVID-19 HGI data freeze 5 (January 18, 2021, https://www.covid19hg.org/). These studies include four phenotypes: 1) A2 = Very severe respiratory confirmed COVID-19 vs. population; 2) B1 = Hospitalized vs. non-hospitalized COVID-19 patients; 3) B2 = Hospitalized COVID-19 patients vs. population; and 4) C2 = COVID-19 patients vs. population. For each phenotype, four GWASs were performed, including two on individuals from European descent and two on multiple ethnicities. All the studies from three phenotypes (A2, B2 and C2) include more than 1 million controls, whereas one (B1), which tested hospitalized and non-hospitalized patients, included < 6,000 cases and < 16,000 controls.

For each study, we identified all variants with p-value < 10^−7^ (considered as “genome-wide-significant”) from the filtered GWAS summary statistics (p < 1 × 10^−5^) provided by the COVID-19 HGI (https://www.covid19hg.org/results/r5/) and expanded each variant’s position by 500 kb upstream and downstream. We used *bedtools merge* ^56^ to identify 24 genome-wide-significant unique loci.

### GTEx data

We downloaded all SNP-gene association tests (including non-significant test) in all GTEx V.7 tissues and gene-level information from the GTEx Portal (https://www.gtexportal.org/home/datasets). In each tissue, we extracted the eQTL information for 679 genes that overlapped the 24 COVID-19 genome-wide significant loci.

### Colocalization

We performed colocalization using two datasets: 1) between each pair of COVID-19 GWAS studies obtained from COVID-19 HGI; and 2) for each gene in the 24 COVID-19 genome-wide significant loci, between its eQTL signal in each of 48 GTEx tissues and each COVID-19 GWAS study. We used the *coloc*.*abf* function from the *coloc* package in R ^14^ to compare the p-values between each pair of SNPs genotyped in both datasets. We considered as each pair of signals “colocalized” if their PP-H4 was greater than 0.9.

### LD calculation

To obtain LD information for each of the most likely causal variants in the colocalization between GWAS and eQTLs (Figure 5), we used LDlinkR ^57^. We obtained a token from https://ldlink.nci.nih.gov/?tab=apiaccess and interrogated the 1000 Genomes LD structure using the *LDproxy* function.

## Data Availability

All data used in this manuscript is publicly available.

## Author information

KAF and MD conceived the study. MD, ADC, TAD, JPN and HM conducted analyses. MD and KAF prepared the manuscript.

## Acknowledgements

This work was supported by Emergency COVID-19 Seed Award from the Tobacco-Related Disease Research Program of the University of California, Grant R00RG2716. JPN and TDA were supported by an NIH training grant (T15LM011271).

## Supplementary information

**Table S1: Genome-wide-significant COVID-19 loci**

The table shows the coordinates of each the 24 loci with a minimum p-value < 10^−7^ in at least one of the 16 GWAS obtained from the COVID-19 HGI data freeze 5 (Table 1).

**Table S2: Colocalization between GWAS signals at each of the 24 genome-wide significant loci**

The table shows pairwise colocalization results of the 16 COVID-19 GWAS at each of the 24 genome-wide significant loci. Shown are: each locus coordinates (chromosome, start and end positions), the two GWAS, the SNP ID of the variant with the highest posterior probability of being causal, the p-values of this variant in both studies, the number of SNPs tested for colocalization, the posterior probabilities of the five colocalization hypotheses; and posterior probability of the lead SNP.

The colocalization hypotheses are: 1) H0: neither study has a significant association at the tested locus; 2) H1: only the first study is associated; 3) H2: only the second study is associated; 4) H3: both studies are associated but the underlying variants are different; and 5) H4: both studies are associated and share the same underlying variants.

**Table S3: Colocalization between GWAS signals and eQTLs in 48 tissues at each of the 24 genome-wide significant loci**

The table shows the pairwise colocalization results between eQTLs and COVID-19 GWAS signals for each gene mapping into the 24 COVID-19 genome-wide significant loci. Shown are: each locus coordinates (chromosome, start and end positions), gene ID, gene name, tissue tested for eQTLs, GWAS study, the SNP ID of the variant with the highest posterior probability of being causal, the p-values of this variant in both studies, the number of SNPs tested for colocalization, the posterior probabilities of the five colocalization hypotheses; and posterior probability of the lead SNP.

